# Delta Variant SARS-CoV-2 infections in pediatric cases during the second wave in India

**DOI:** 10.1101/2021.12.09.21266954

**Authors:** Pragya D. Yadav, Gunjan Kumar, Aparna Mukherjee, Dimpal A. Nyayanit, Anita M. Shete, Rima R. Sahay, Abhinendra Kumar, Triparna Majumdar, Savita Patil, Priyanka Pandit, Yash Joshi, Manisha Dudhmal, Samiran Panda, Lokesh Kumar Sharma, ML Kala Yadav, Jayanthi Shastri, Mayank Gangwar, Ashok Munivenkattapa, Varsha Potdar, K Nagamani, Kapil Goyal, Ravisekhar Gadepalli, Maria Thomas, Suruchi Shukla, P Nagraj, Vivek Gupta, Gaurav Dalela, Nawaz Umar, Sweety M Patel

**Author notes:** **Correspondence:** Maximum Containment Laboratory, Indian Council of Medical Research-National Institute of Virology, Sus Road, Pashan, Pune 411021, India /290. equal first author (Pragya D. Yadav, Gunjan Kumar, Aparna Mukherjee). All authors have contributed equally (ML Kala Yadav, Jayanthi Shastri, Mayank Gangwar, Ashok Munivenkattapa, K Nagamani, Kapil Goyal, Ravisekhar Gadepalli, Maria Thomas, Suruchi Shukla, P Nagraj, Vivek Gupta, Gaurav Dalela, Nawaz Umar, Sweety M Patel).

## Abstract

The aim of this study was to identify the SARS-CoV-2 lineages circulating in the pediatric population of India during the second wave of the pandemic. Clinical and demographic details linked with the nasopharyngeal/oropharyngeal swabs (NPS/OPS) collected from SARS-CoV-2 cases (n=583) aged 0-18 year and tested positive by real-time RT-PCR were retrieved from March to June 2021.Symptoms were reported among 37.2% of patients and 14.8% reported to be hospitalized. The E gene CT value had significant statistical difference at the point of sample collection when compared to that observed in the sequencing laboratory. Out of these 512 sequences 372 were VOCs, 51 were VOIs. Most common lineages observed were Delta, followed by Kappa, Alpha and B.1.36, seen in 65.82%, 9.96%, 6.83% and 4.68%, respectively in the study population. Overall, it was observed that Delta strain was the leading cause of SARS-CoV-2 infection in Indian children during the second wave of the pandemic. We emphasize on the need of continuous genomic surveillance in SARS-CoV-2 infection even amongst children.

## Introduction

The first case of the severe acute respiratory syndrome coronavirus −2 (SARS-CoV-2) was reported from Wuhan, China during December 2019 (1). In India, the first wave of the SARS-CoV-2 pandemic started in March 2020, which peaked in mid-September 2020 infecting approximately 5.7 million population(2). The predominant SARS-CoV-2 variant leading to coronavirus infection-19 (COVID-19) had a D614G mutation in the spike protein as compared to the parent strain, reported from Wuhan, China. The D614G mutation at the spike protein region was reported to have increased transmissibility (3).

Newer variants of SARS-CoV-2, having amino acid mutation in the spike protein, were reported from different countries from December 2020 that had increased ability for transmission and evasion of human immune response in comparison to earlier strains. The 20I/S: 501Y.V1 (currently alpha variant; Pango lineage: B.1.1.7) was the earliest to be reported from United Kingdom (UK) during December 2020 (4). Emergence of variants with mutations across different genes in the SARS-CoV-2 genome led to outbreaks in different countries similar to UK. The Center for Disease Control and Prevention, USA (CDC) classified the virus based on transmissibility, virulence and diagnostic availability in three classes; a) variant of concern (VOC), b) variant of interest (VOI) and c) variant of high consequences (5). The rise in the SARS-CoV-2 variants led to the development of Phylogenetic Assignment of Named Global Outbreak LINeages (PANGOLIN) classification, which is currently being used for identification of new SARS-Cov-2 variants. The World Health Organization (WHO) has labeled these variants using the Greek alphabets (alpha, beta, gamma, delta and so on) based on their reporting dates (6). Till date SARS-CoV-2 variants circulating in India were reported belonging to the pangolin lineage of B.1.1.7 (Alpha), B.1.351 (Beta) and B.1.1.28.1 (Gamma), B.1.1.28.2 (Zeta), B.1.617.1 (Kappa), B.1.617.2 (Delta) and Delta derivatives as well as B.1.617.3 (7–9).

There is limited information pertaining to the SARS-CoV-2 strain infecting the children in our country. A systematic review of the published literature between January-March 2020 demonstrated less than 5% of children to be SARS-CoV-2 positive, with mild clinical symptoms as compared to adults (10). Yonker et al showed the detection of SARS-CoV-2 variants belonging to the Alpha and B.1.526.2 lineages (derivatives of Iota), identified in the age group below 21 years (11).

The current study was performed to identify the SARS-CoV-2 lineages circulating in the pediatric population across 8 States and one Union Territory in India during the second wave of the pandemic during March to June which attained its peak in May 2021. Additionally, the Indian pediatric SARS-CoV-2 sequences available with GISAID database with collection date from January 2020 to July 31, 2021were also analyzed.

## Materials and Methods

### Study Sites and retrieval of the clinical data

The National Clinical Registry for COVID-19 is a data collection platform for hospitalized COVID-19 patients maintained by the Indian Council of Medical Research (ICMR) in collaboration with the Ministry of Health & Family Welfare (MOHFW), All India Institute of Medical Sciences (AIIMS), New Delhi and ICMR-National Institute of Medical Statistics (NIMS). The structure and protocol of the registry are available in the public domain. (https://www.icmr.gov.in/tab1ar1.html). Currently, forty one tertiary care hospitals, both public and private, across India, are involved in this registry. All participating hospitals were invited to send stored nasopharyngeal samples of COVID-19 positive children aged 0-18 year. Twelve participating institutes and in addition two VRDL laboratories agreed to send samples. These collections were carried out by designated laboratories. Clinical and demographic details of SARS-CoV-2 cases (n=583) aged 0-18 year, tested positive by real-time RT-PCR were retrieved using the central testing database from ICMR. The samples collected had representation from 8 States and one Union Territory (Chandigarh), and were collected during a period of March 2020 to June 2021. (Maharashtra, Gujarat, Uttar Pradesh, Karnataka, Rajasthan, Madhya Pradesh, Punjab and Andhra Pradesh) (www.icmr.gov.in).

### Inclusion, exclusion criteria and transport of specimens

SARS-CoV-2 cases fulfilling the following inclusion criteria were enrolled under the study: i) Nasopharyngeal/Oropharyngeal (NPS/OPS) swab samples collected fromSARS-CoV-2 cases during the period of second wave ii) Cases whose real time RT-PCR threshold value was <30 and NPS/OPS were appropriately stored at −80 °C; (iii) Sample referral forms (SRF) capturing the demographic and clinical details of cases were available with the respective sites and (iv) Provision of the consent from the parents/guardians. The NPS/OPS samples of all these cases which fulfilled all the inclusion criteria were packed in triple-layer packaging with dry ice as per International Air Transport Association (IATA) guidelines and then transferred to the Reference laboratory the ICMR-National Institute of Virology (ICMR-NIV), Pune for next generation sequencing (NGS) and variant analysis.

### RNA extraction and NGS on the clinical samples

Total RNA was extracted from nasal or throat swab specimens using the Magmax RNA extraction kit(Thermofisher, USA). Extracted RNA was checked by SARS CoV-2 specific real time RT PCR. For preparation of RNA libraries Illumina CovidSeq protocol (Illumina Inc, USA) was followed (12). In brief, the first strand synthesized from extracted RNA was amplified using two primers pools followed by tagmentation. This tagmented amplicon was further used for the preparation of libraries as per manufacturer’s instructions. The libraries were pooled, purified, quantified and then loaded at final loading concentration of 1.4picomole onto the NextSeq 500/550 system using NextSeq 500/550 High Output Kit v2.5 (75 Cycles) (Illumina Inc).

The paired-end FASTQ files generated were analyzed on the CLC Genomics Workbench version 21.0.4 (CLC, Qiagen). Wuhan Hu-1 isolate (Accession Number: NC_045512.2) was used for reference based assembly to retrieve the genomic sequence of the SARS-CoV-2. The retrieved sequences were deposited in the public repository of Global Initiative on Sharing Avian Influenza Data (GISAID) database to make the data available for others.

### Phylogenetic and sequence analysis

The Indian pediatric SARS-CoV-2 sequences having collection date from January 2020 to July 31, 2021 were downloaded (n=1751) from the GISAID database (13). Representative sequences based on the data from the different States/Union Territory and the specific clades were used in the analysis along with the sequences retrieved in this study. The sequences were aligned using the CLC genomics workbench. The aligned file was manually checked for correctness. A neighbor-joining phylogenetic tree was constructed from the coding region of the SARS-CoV-2 genome using the maximum composite likelihood model along with gamma distribution as the rate variation parameter. A bootstrap replication of 1000 cycles was performed to assess the statistical robustness of the generated tree. The amino acid variation for each gene and net nucleotide and amino acid divergence was identified using the MEGA software version 7.0(14) and illustrated using GraphPad Prism v9.

### Statistical Analysis

Data analysis was done using STATA v14 (College Station, TX). The continuous variables were described as mean with standard deviation (SD) or median with inter quartile range (IQR). The categorical variables were described as frequency and proportions. Association between categorical variables was tested by applying chi square test. Difference amongst the continuous variables was tested using paired and unpaired student’s t-test or ANOVA as required. P value less than 0.05 was considered to be significant at 95% confidence interval (CI).

## Results

### Clinico-demographic analysis

We present here the results of 583 samples included in the current study. Table 1 shows the demographic and clinical characteristics of the enrolled children at baseline. More than half the samples were from males and the median (IQR) age of the study participants was 13 year (8, 16).More than half of the patients (51.8%) belonged to the age group of 13-19 year, 41.2% to 3-12 year and the rest 7% to less than 3 year. Symptoms were reported among 37.2% of patients and 14.8% reported to be hospitalized. Symptom profile was available for only 74 patients, with fever, cough, runny nose and sore throat, being the most common symptoms noted in 60%, 49.3%, 23.4% and 12% of the children, respectively. The rest of the symptoms were seen in less than 10% of the patients. Significant statistical difference was observed in the E gene CT value at the point of sample collection and at ICMR-NIV, Pune, with that being done at ICMR-NIV to be lower. The CT values were not significantly different among the lineages, among males and females, different age groups, hospitalized and non-hospitalized patients, though it was significantly lower among symptomatic and hospitalized patients [Table-2].

**Table 1:** Characteristics of the study participants at the time of sample collection, n=583.

| Characteristic | Values |
| --- | --- |
| Age in year, Median (IQR) | 13 (8,16) |
| **Age categories** |  |
| 0-2 year | 41 (7) |
| 3-12 year | 240 (41.2) |
| 13-18 year | 302 (51.8) |
| Boys | 319 (55.3) |
| Symptomatic, n=554 | 206 (37.2) |
| Hospitalized, n=501 | 74 (14.8) |
| **Symptom profile, n=74** |  |
| Fever | 42 (60) |
| Cough | 35 (49.3) |
| Runny Nose | 15 (23.4) |
| Sore Throat | 8 (12.5) |
| Body ache | 5 (7.8) |
| Breathlessness | 4 (6.3) |
| Vomiting | 3 (4.7) |
| Fatigue | 1 (1.6) |
| Eye pain | 1 (1.6) |
| **Centre wise samples** |  |
| Bowringand lady Curzon Medical College, Bangalore, Karnataka | 203 (34.8) |
| Kasturba Hospital of Infectious diseases, Mumbai, Maharashtra | 69 (11.8) |
| Institute of Medical Sciences, Banaras Hindu University, Varanasi, Uttar Pradesh | 46 (7.9) |
| National Institute of Virology, Bangalore Unit | 38 (6.5) |
| Gandhi Hospital, Secunderabad, Telangana | 36 (6.2) |
| Postgraduate Institute of Medical Education &Research, Chandigarh | 36 (6.2) |
| All India Institute of Medical Sciences, Jodhpur, Rajasthan | 33 (5.7) |
| Christian Medical College, Ludhiana, Punjab | 28 (4.8) |
| King George Medical University, Lucknow, Uttar Pradesh | 28 (4.8) |
| Gandhi Medical College, Bhopal, Madhya Pradesh | 20 (3.4) |
| Government Institute of Medical Sciences, Noida, Uttar Pradesh | 16 (2.7) |
| Sawai Man Singh Medical College, Jaipur, Rajasthan | 14 (2.4) |
| Gulbarga Institute of Medical Sciences, Gulbarga, Karnataka | 9 (1.5) |
| Smt. NHL Municipal Medical College, Ahmedabad, Gujarat | 7 (1.2) |

<sup>1</sup>Values are expressed as n (%) unless specified.

**Table 2.**
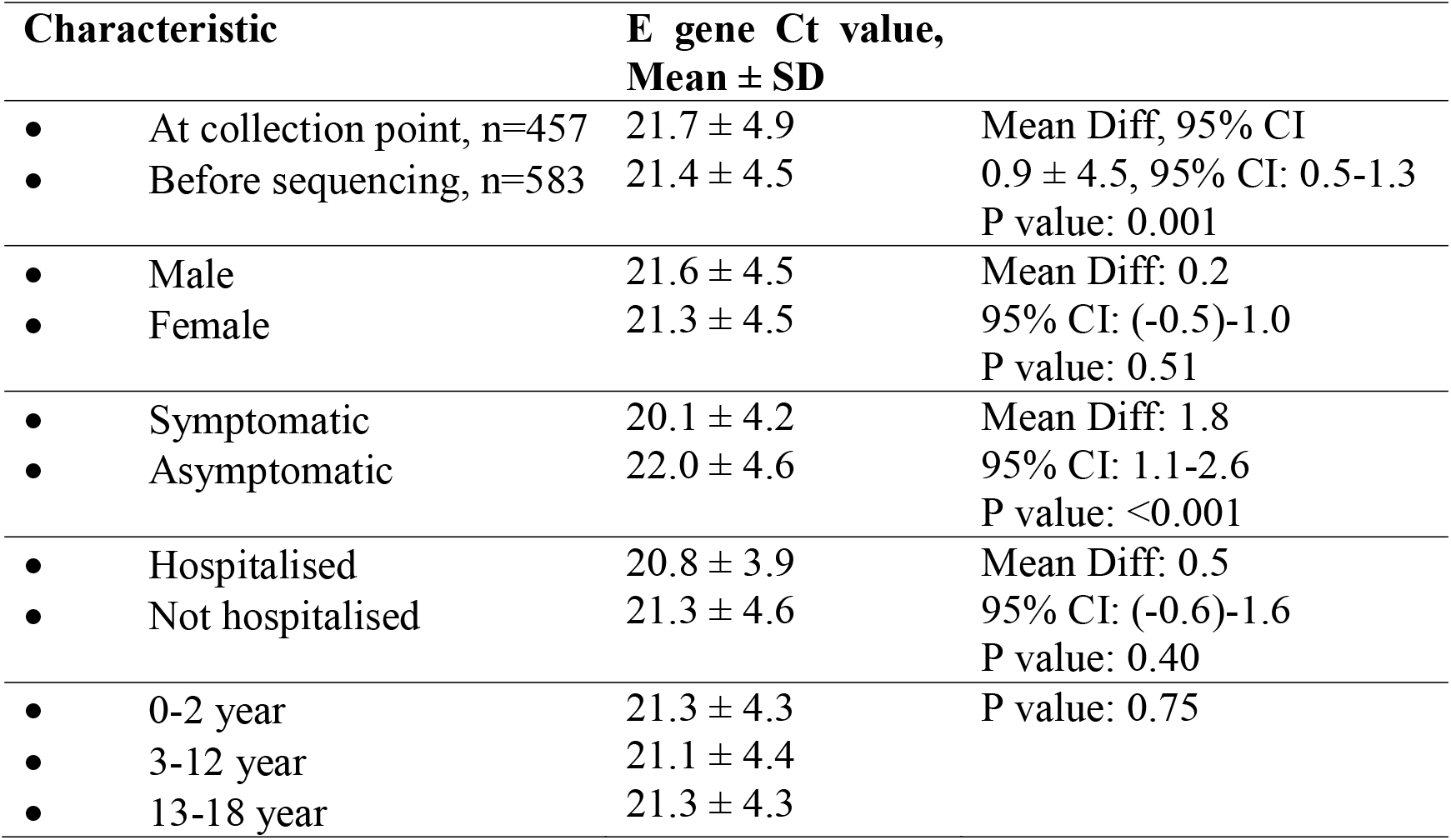
Ct values among the study participants by grouping in different categories.

### SARS-CoV-2 data analysis, NGS and phylogenetic analysis

In order to understand the dominance of SARS-CoV-2 variant present during the first wave of its outbreak, the 1751 pediatric SARS-CoV-2 sequences were grouped into two based on the presence of D614G mutation in the spike region. The month-wise observation of the groups is depicted in Figure-1. Month-wise cases reported without D614G mutations revealed the presence of early GISAID clades (L, S, and V) (Figure-1A). The predominance of these variants was observed till August 2020 in pediatric sequences (Figure-1A). The first SARS-CoV-2 to be reported belonged to unclassified cluster (‘O’) with few cases of the other clades [L, S, and V]. Month-wise cases reported with D614G mutation (‘G’ clade) is depicted in Fig 1B.TheG clade variant of SARS-CoV-2 was reported as early as March 2020. This variant along with its sister clades predominated until February 2021, the first wave of SARS-CoV-2 in India. The amino acid changes observed with the sister clades are GH(S: D614G, NS3: Q57H; Greek name: Beta) and GR(S: D614G, N: G204R; Greek name: Gamma).

**Figure 1.**
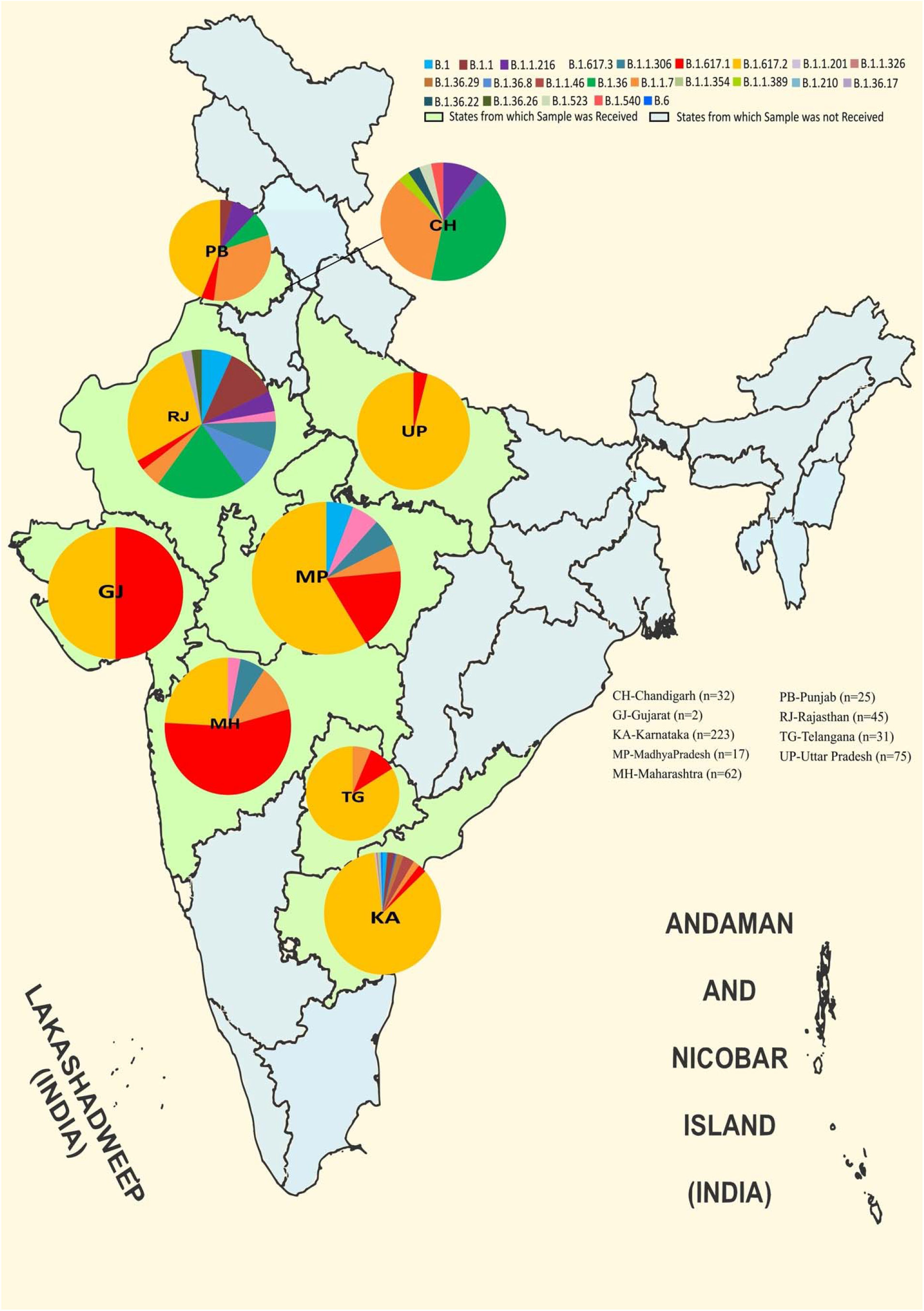
Month wise distribution of the GISIAD clade: A month –wise distribution of pangolin lineages classified under A) without D614G and B) with D614G mutation. The x-axis depicts the different months in year 2020-2021 and the Y-axis is the Log2 scale of SARS-CoV-2 cases reported in GISAID.

As observed in the adult population, the new SARS-CoV-2 variants were also reported in the pediatric cases from India. These variants were GK(S-D614G + S-T478K; Greek name: Delta), GRY (S-H69del, S-V70del, S-Y144del, S-N501Y + S-D614G + N-G204R; Greek name: Alpha) and GV(S-D614G + S-A222V). The GRY variant was the first to be reported, beginning from November 2020 followed by the GK variant from December 2020. The reporting of these variants increased rapidly during the consequent months. They superseded earlier GR (Gamma) and GH (Beta) variant of SARS-CoV-2 during March 2021 which also marks the beginning of SARS-CoV-2’s second wave in India (Figure-1B).The B.1.617.2 (Delta) variant showed a steep rise after March 2021, reaching to 100% amongst the collected samples in June. (Figure-1B).

Out of the 583 samples that were used for NGS, SARS-CoV-2 sequences with 98.5% genome coverage were retrieved from 512 samples. The details of the percent genome coverage, relevant read mapped and the total reads for each sample along with their EPI-Accession numbers are given in Supplementary Table-S1. These sequences were classified into PANGOLIN lineage using https://pangolin.cog-uk.io/. The PANGOIN lineage of the SARS-CoV-2 sequences is also given in Supplementary Table 1. Out of these 512 sequences 372 were VOCs, 51 were VOIs and 89 were the other reported variants. Most common lineages observed were Delta, followed by Kappa, Alpha and B.1.36, seen in 65.82%, 9.96%, 6.83% and 4.68%, respectively in the study group.

Out of 1752 SARS-CoV-2 sequences from pediatric age group, 1609 sequences were used for further analysis based on the complete genome coverage and high-quality reads. A total of 2121 SARS-CoV-2 sequences [comprising of 1609 sequences from GISAID database and 512 SARS-CoV-2 sequences retrieved from this study] were analyzed further. It was observed that the Kappa was first reported in the pediatric case as early as September 2020 followed by Delta during October 2020 (Figure-2A). It is notable that no VOCs and VOIs were reported in the pediatric population prior to September 2020. The Alpha variant was reported later to Kappa from December 2020, which is reversed in case of adult population.

**Figure 2.**
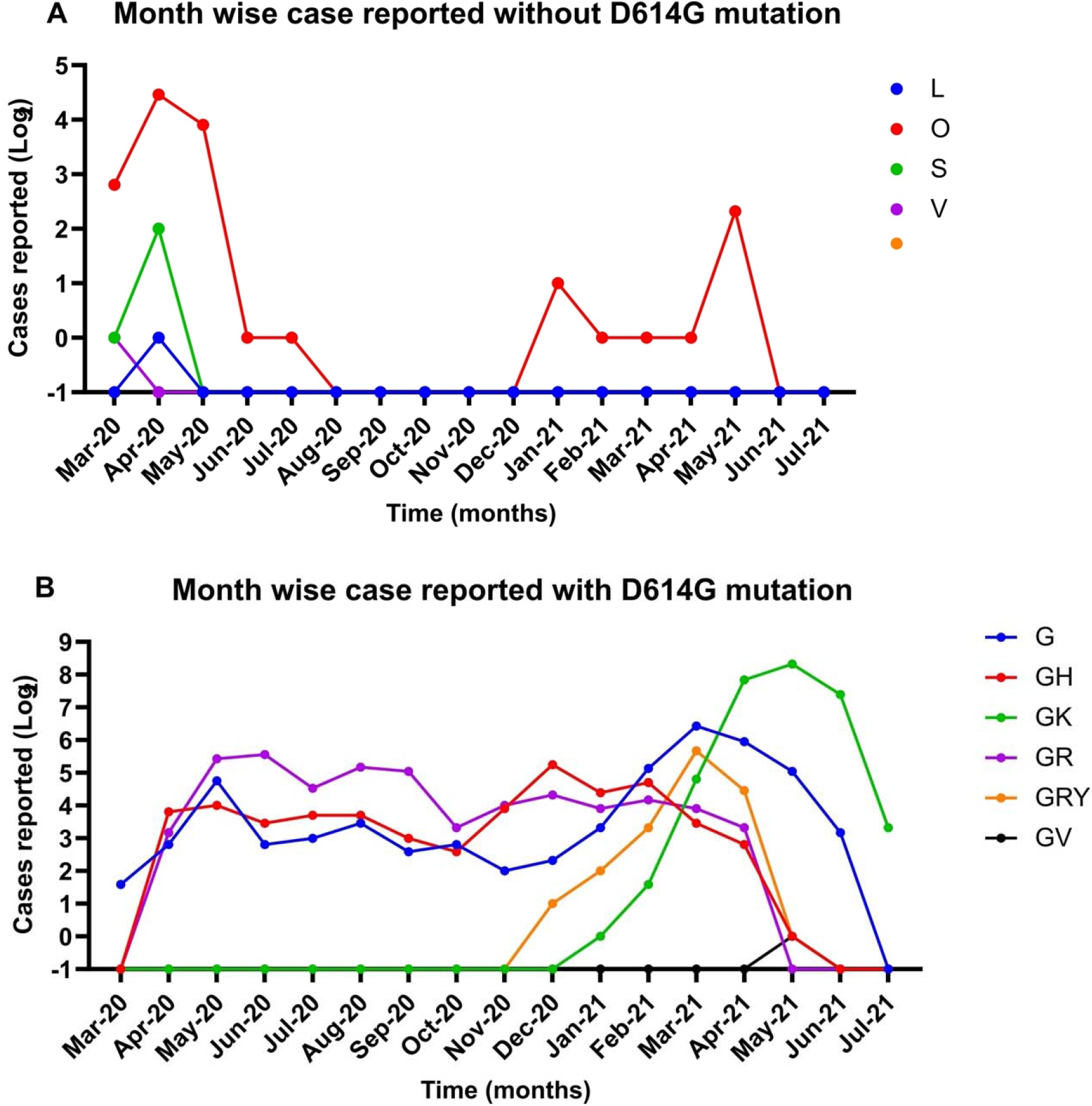
Month wise distribution of the Pangolin lineages: A month –wise distribution pangolin lineages classified under A) VOC and VOI and B) other lineages. The x-axis depicts the different months in year 2020-2021 and the Y-axis is the Log2 scale of SARS-CoV-2 cases reported in GISAID and retrieved during this study.

During January-March of 2021, a higher number of Alpha variant cases were reported, which was superseded rapidly by Delta (Figure-2A) amongst children. The Delta variant is currently also the most predominant strain circulating amongst pediatric population. Along with these a few other VOCs and VOIs were also reported (Figure-2B).

The lineage distribution of the 512 SARS-CoV-2 sequences retrieved across the different states studied is depicted in Figure-3. It was observed that Delta lineage predominated in Karnataka, Uttar Pradesh, Madhya Pradesh and Telangana whereas the Kappa variant was observed in Maharashtra, Rajasthan and Chandigarh. The northern part of India revealed the presence of SARS-CoV-2 strains apart from the VOC and VUIs. Overall, it was observed that Delta strain predominated as the leading cause to SARS-CoV-2 infection in children in India during the second wave of the pandemic.

**Figure 3.**
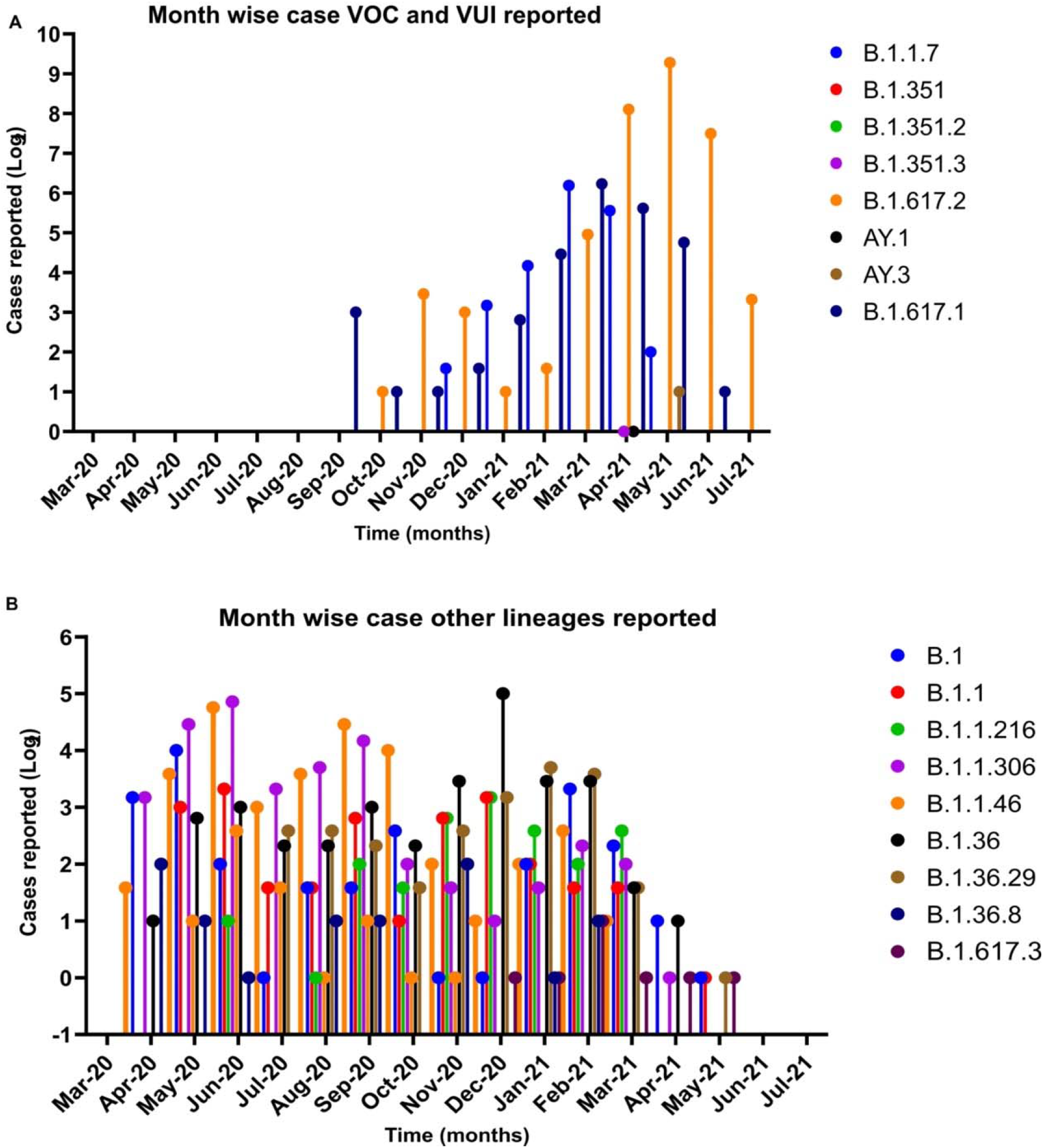
Distribution of the SARS-CoV-2 genome prevalence among cases pediatric cases in India. The distribution in the pie chart is proportional to the numbers in each respective clade in each state. The outline of India’s map was downloaded from http://www.surveyofindia.gov.in/file/Map%20f%20India_1.jpg (accessed on 20 March 2020) and further modified to include relevant data in the SVG editor.

The maximum likelihood tree was generated using the GTR+G as substitution model and a bootstrap replication of 1000 cycles. A separate cluster of B.1.617 lineage was observed that segregated into Kappa, Delta and B.1.617.3 lineages as depicted in Figure 4.

**Figure 4:**
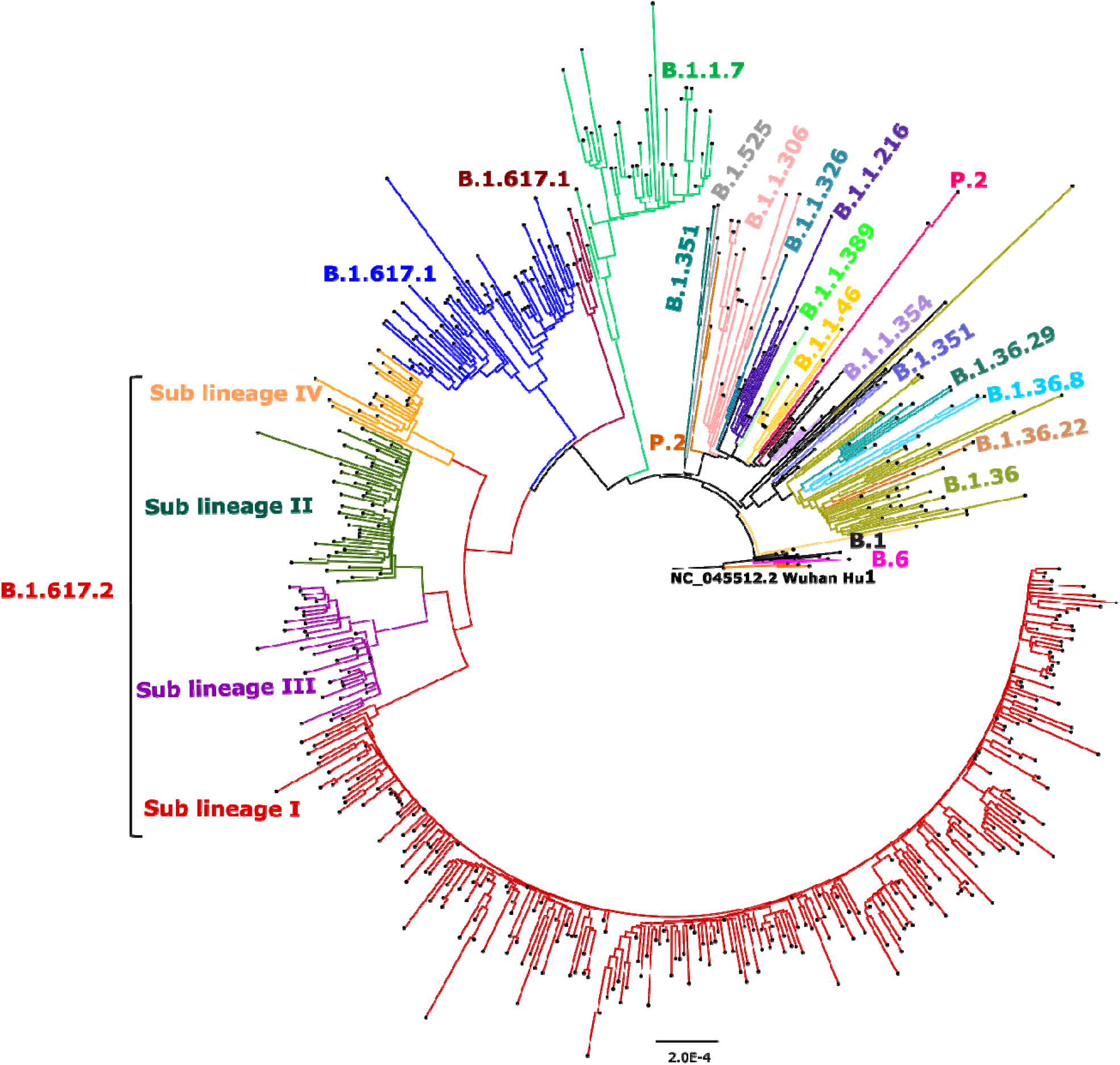
Maximum likelihood tree for the SARS-CoV-2 genomes retrieved from pediatric cases. Representative SARS-CoV-2 sequences from different lineages along with 512 sequences retrieved were used to generate the tree with a bootstrap replication of 1000 cycles. The sub-lineages I–IV of B.1.617.2are marked in red, green, pink, and orange color on the nodes, respectively. B.1.617.1 sequence is marked in brown and B.1.617.3 in blue color. The other retrieved pangolin lineages are also marked on branches in different colors. Fig Tree v1.4.4 and Inkscape were used to visualize and edit the generated tree.

## Discussion

India after the completion of massive immunization of 1 billion doses of COVID-19 in the adult population has recently approved two vaccines for pediatric use namely Covaxin/BBV152 (inactivated virus vaccine) and ZyCoV-D (DNA vaccine), both vaccines are being manufactured in India. Advocacy for re-opening of schools based on evidence was ongoing (15) and many states have started taking cognizance of the same. The plausibility of a third wave in India has also been extensively studied and helped programmatic discussion and decision making(16). Studies underline that children aged 1-17 year are susceptible to a milder form of COVID-19 but not to severe disease in comparison to the adult population (17). The reasons behind this were mainly the lower density of angiotensin-converting enzyme-2 (ACE2) receptors lining the respiratory tract (17). The 4th serosurvey of COVID-19 in India showed that more than 50%children belonging to 6-17 year age group were seropositive emphasizing that a huge number of children were infected as adults (18,19), but not many of them required institutional care.

Genome sequencing of SARS-CoV-2 has played an important role in determining its emerging variants, geographical predominance, evolution over different times and understanding its trend. In India during the first wave of SARS-CoV-2 the variant with spike protein D614G mutation had dominated. The epidemiological SARS-CoV-2 analysis of adult population from India demonstrated the GH and GR clades to predominate in the northern and southern part of India respectively (20,21). The current study is a molecular epidemiology study which explored the SARS-CoV-2 distribution in the pediatric cases from different parts of India during the first and second wave of pandemic. It was observed that the majority of the pediatric cases reported during the first wave did not have the spike protein D614G mutation and later B.1.617 lineage was found to be predominant in children as observed in adult SARS-CoV-2 infections(20). Further the four derivatives of Delta were also observed in the pediatric sequences as described earlier (22,23). It was observed that sub-lineage I had majority of the Delta lineage shared followed by sub lineage-II (Figure-4).

The second wave of the COVID-19 in the world witnessed the emergence of variants of concern (Alpha, Beta, Gamma, Delta) and VOIs (Kappa, Zeta, Lambda (C.37)] in the adults. In India, the first pediatric case infected with Alpha strain was reported in November 2020.Further an increase in the number of cases was observed with Alpha and the Delta variants. It was observed that majority of the pediatric cases across the states surveyed in the current study had Delta variant indicating it to be a predominant strain with amino acid changes in ORF1ab (A1306S, P2046L, P2287S, V2930L, T3255I, T3446A, G5063S, P5401L, A6319V) and N G215C.This study identifies the SARS-CoV-2 variant responsible for the infection in the pediatric population and highlights the importance of genomic surveillance in children.

## Data Availability

All data produced in the present study are available upon reasonable request to the authors

## Author Contributions

***Conception and design of study***: P D. Yadav, A Mukherjee, S Panda ***Acquisition of data (laboratory or clinical***): P D. Yadav, S Panda, G Kumar, AMukherjee, K Yadav ML, J Shastri, M Gangwar, A Munivenkattapa, V Potdar, K Nagamani, K Goyal, R Gadepalli, M Thomas, S Shukla, Nagraj, V Gupta, G Dalela, N Umar, S M Patel, L K Sharma, R R. Sahay, A Kumar, D A. Nyayanit, A M. Shete, S Patil, T Majumdar, M Dudhmal, P Pandit and Y Joshi ***Data analysis and/or interpretation*:** P D. Yadav, D A. Nyayanit, G Kumar, A Mukherjee ***Drafting of manuscript and/or critical revision*:** P D. Yadav, G Kumar, D A. Nyayanit, A Mukherjee ***Approval of final version of manuscript*:** P D. Yadav, A Mukherjee, S Panda, G Kumar, K Yadav ML, J Shastri, M Gangwar, A Munivenkattapa, V Potdar, K Nagamani, K Goyal, R Gadepalli, M Thomas, S Shukla, Nagraj, V Gupta, G Dalela, N Umar, S M Patel, L K Sharma, R R. Sahay, A Kumar, D A. Nyayanit, A M. Shete, S Patil, T Majumdar, M Dudhmal, P Pandit and Y Joshi

## Institutional Review Board Statement

The study was approved by the Institutional Human Ethics Committee of ICMR-NIV, Pune, India under project ‘Molecular epidemiological analysis of SARS-CoV-2 circulating in different regions of India’(IHEC no. NIV/IEC/Dec/2020/D-6 dated 31 December 2020).

## Informed Consent Statement

Patient consent was waived due to screening of retrospective samples (TS/NS) already collected during Covid-19 pandemic by all the respective centers involved in the study.

## Data Availability Statement

All the sequencing data and information of this study is available in GISAID. Accession no is provided in supplementary table.

## Acknowledgments

Authors gratefully acknowledge the encouragement and support extended by Dr. Balram Bhargava, Secretary to the Government of India, Department of Health Research, Ministry of Health and Family Welfare and Director-General, ICMR and Prof. (Dr.) Priya Abraham, Director, ICMR-NIV, Pune. We are grateful to Mrs. Ashwini Waghmare, Ms. Pranita Gawande, Mrs. Kaumudi Kalele, Ms. Jyoti Yemul and Mrs. Tejashri Kore for excellent technical support during the study. We thank to all contributors those have submitted the SARS-CoV-2 sequences in the GISAID database

## Supplementary Materials

The details of the percent genome coverage, relevant read mapped, the total reads and the EPI-Accession numbers for samples studied.

